# Influenza activity in the United States during the 2025-26 influenza season

**DOI:** 10.64898/2026.11.16.26367927

**Authors:** Alexandra L. Busbee, Amanda C. Howa, Yingtao Zhou, Raheem Smith, Nicholas Dempster, Peter Daly, Angiezel Merced-Morales, Arielle Colon, Daneisha Hawkins, Shunte Moon, Katie Reinhart, Stacy Huang, Chao Zhang, A. Danielle Iuliano, Alissa O’Halloran, Jenn Habeck, Dawud Ujamaa, Catherine H. Bozio, Sarabeth Mathis, Ha T. Nguyen, Mira C. Patel, Larisa Gubareva, Xiao-Yu Zheng, Rebecca Kondor, Krista Kniss, Alicia Budd

## Abstract

**Background:** Comprehensive influenza surveillance is conducted year-round in the United States to monitor changes in influenza activity. This report summarizes the 2025-26 U.S. influenza season.

**Methods:** Data from September 28, 2025 through October 3, 2026 were collected from national surveillance systems monitoring influenza through laboratory testing, outpatient and emergency department visits, hospitalizations, and mortality. Data modeled disease burden and assessed severity using established criteria. A subset of influenza positive specimens were genetically and antigenically characterized and assessed for antiviral susceptibility.

**Results:** Influenza activity peaked during the week ending December 27, 2025, five weeks earlier than the 2024-25 season. There were two waves of influenza activity with influenza A(H3N2) subclade K viruses predominating the season overall and during the first wave followed by a smaller B/Victoria wave. The majority of circulating A(H3N2) and B/Victoria viruses were antigenically drifted from 2025-26 influenza vaccine reference strains.

Season severity was classified as moderate overall, with children (0-17 years old) experiencing a high severity season and adults experiencing a moderate severity season. Children experienced the highest percent of influenza-associated outpatient and emergency department visits compared to other age groups. This season’s cumulative influenza-associated hospitalization rate in children is the highest hospitalization rate in the past 16 years. Additionally, there were 198 influenza-associated pediatric deaths reported to CDC as of October 29, 2026.

**Conclusions:** Although antigenically drifted A(H3N2) viruses predominated, the 2025-26 U.S. influenza season occurred within historic norms for timing and severity. Year-round surveillance remains critical for detecting changes in influenza viruses and monitoring influenza-associated morbidity and mortality.

**Key Points:**

- CDC’s severity assessment framework classified the 2025-26 U.S. influenza season as moderate across all age groups
- Nationally, peak influenza activity occurred in late December 2025
- Influenza A(H3N2) subclade K viruses predominated during the 2025-26 season

## Introduction

In the United States (U.S.), an estimated 8% of the population becomes ill with influenza each year,^1^ although incidence, disease burden, and severity vary seasonally. The Centers for Disease Control and Prevention (CDC) estimates influenza infections result in 9.4 - 51 million illnesses, 120,000 - 710,000 hospitalizations, and 6,300 - 52,000 deaths annually.^2^ While influenza viruses circulate year-round, seasonal influenza activity increases during fall and winter months, with peak timing most often occurring between December and February.^3^

The national influenza surveillance system identifies changes in circulating viruses, detects human infections with novel influenza A viruses, monitors seasonal trends, and measures influenza-associated morbidity and mortality. It is a collaborative surveillance network coordinated by CDC in partnership with state, local, and territorial public health departments; health care providers and facilities; public health and clinical laboratories; and vital statistics offices.^4^

Influenza surveillance data are compiled, analyzed, and reported weekly through CDC’s *FluView* report and online module, *FluView Interactive*.^5,6^ Data are evaluated at the national, Health and Human Services (HHS) regional, and for some systems, state or core based statistical area (CBSA) level.^7,8^ Data are analyzed based on the Morbidity and Mortality Weekly Report (MMWR) week number in which the event (e.g. hospitalization) occurred.^9^

This report provides a comprehensive summary of the 2025–26 influenza season in the U.S., from September 28, 2025 (MMWR Week 40) through week ending October 3, 2026 (MMWR Week 39). Data were reported as of October 29, 2026. This activity was reviewed by CDC, deemed not research, and was conducted consistent with applicable federal law and CDC policy.

### Virus Surveillance

CDC collects influenza virus surveillance data from clinical and public health laboratories.^4^ Clinical laboratories reported results of respiratory specimens tested for influenza as a part of clinical diagnosis. These data were used to track influenza activity, timing, and intensity. Public health laboratories tested influenza viruses to identify influenza A virus subtypes and influenza B virus lineages, and their distribution was monitored throughout the influenza season. Demographic information associated with influenza specimens was reported when available.

During the 2025–26 influenza season, reporting clinical laboratories tested 3,231,299 respiratory specimens for influenza viruses using clinical diagnostic tests. Among these, 329,832 (10.2%) tested positive, including 233,941 (70.9%) for influenza A and 95,891 (29.1%) for influenza B viruses. Nationally, the percent of specimens positive for any influenza virus (all influenza A and B viruses), and for influenza A, peaked at 31.7% and 30.0%, respectively, during the week ending December 27, 2025 (MMWR Week 52), while influenza B peaked at 11.2% during the week ending March 7, 2026 (MMWR Week 9) (**Figure 1**). This overall peak was tied with the 2024–25 overall season peak as the highest percent positive peak since clinical and public health laboratory data were first reported separately in the 2015–16 influenza season (**Supplementary Figure 1**). Most of the U.S. (HHS regions 1-8) peaked in MMWR Week 52, while HHS regions 9 and 10 (West Coast and Pacific Northwest) peaked later in the week ending February 7, 2026 (MMWR Week 5) (**Supplementary Figure 2**).

**Figure 1.**
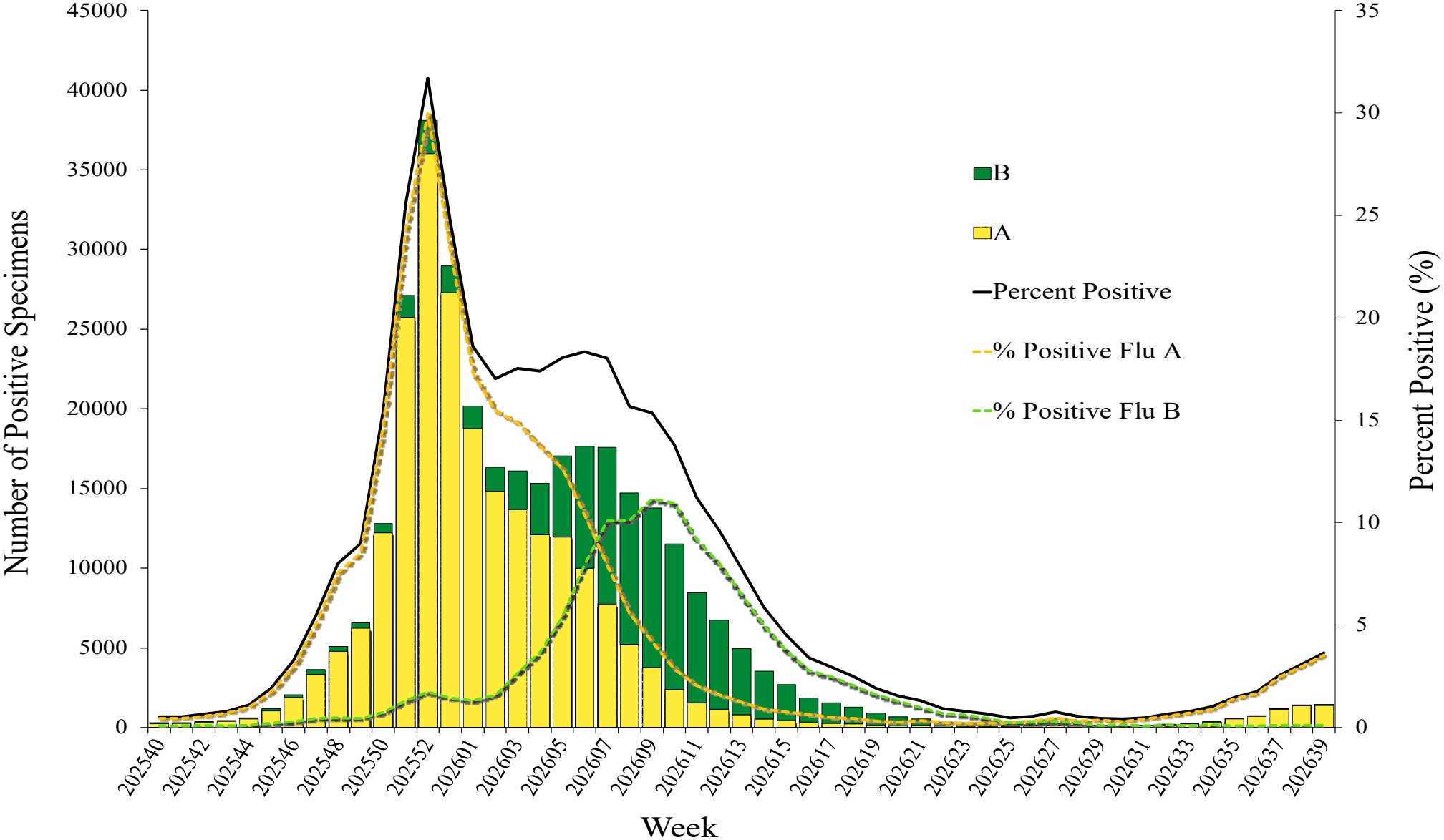
National percent of specimens testing positive for any influenza virus, influenza A, and influenza B and the number of influenza A and B specimens testing positive, by Morbidity and Mortality Weekly Report (MMWR)^a^ Week as reported to the Centers for Disease Control and Prevention (CDC) during the 2025-26 influenza season by a subset of United States clinical laboratories, September 28, 2025 – October 3, 2026 (MMWR Weeks 40-39) ^a^MMWR weeks are defined by the Saturday week-ending date. For influenza surveillance, each season begins in MMWR Week 40, the first week of October, and ends in MMWR Week 39, the last week of September of the following year **Alt text**: Combination bar and line graph showing the number of positive influenza A and influenza B specimens and the percent of positive influenza A and influenza B specimens by MMWR week.

**Figure 2.**
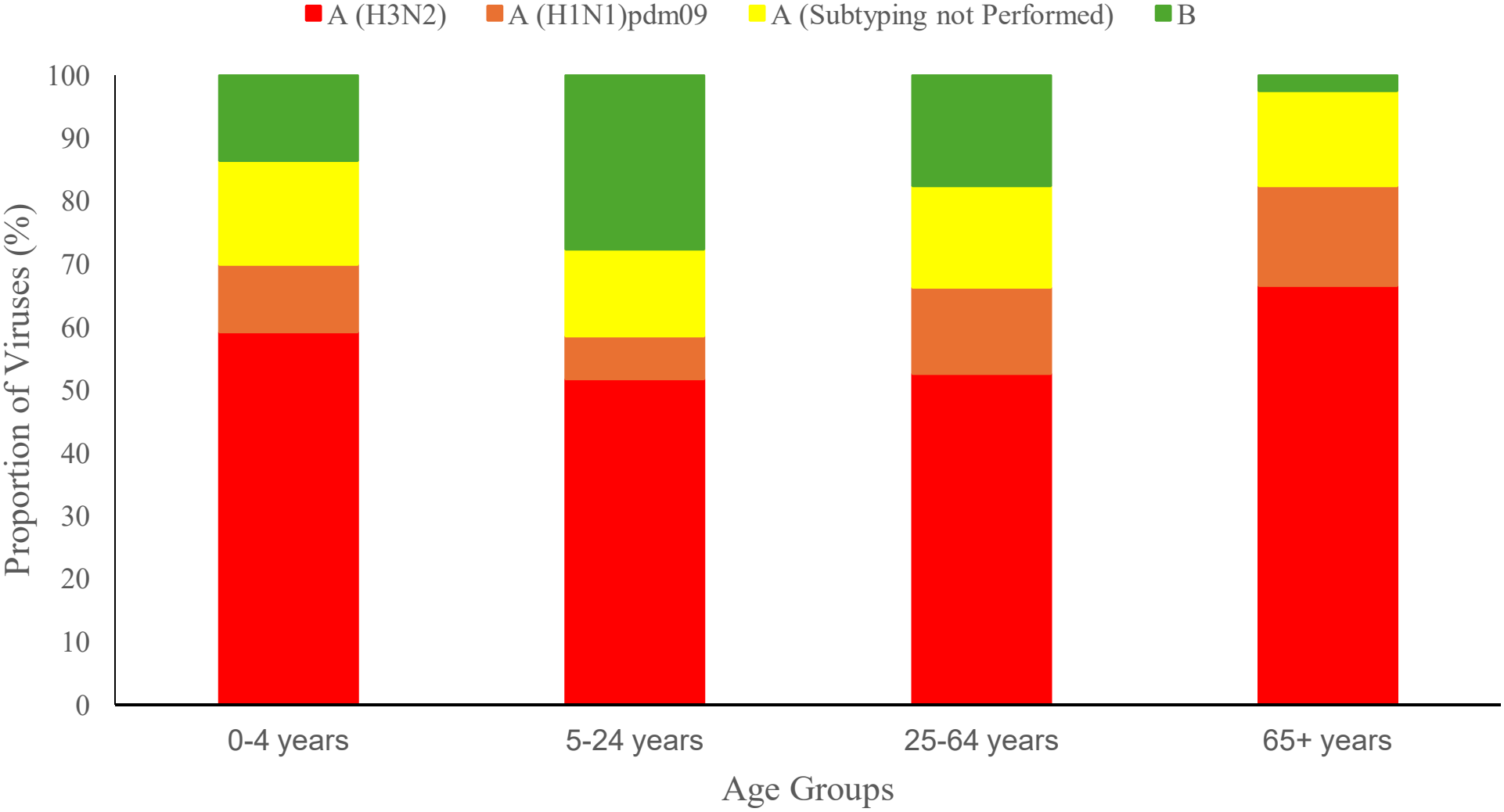
National summary of the proportion (%) of influenza viruses by age group among specimens reported to the Centers for Disease Control and Prevention (CDC) by United States public health laboratories during the 2025-26 influenza season, United States, September 28, 2025– October 3, 2026 (Morbidity and Mortality Weekly Report (MMWR)^a^ Weeks 40-39) *B includes B/Victoria and influenza B viruses with unspecified lineage. ^a^MMWR weeks are defined by the Saturday week-ending date. For influenza surveillance, each season begins in MMWR Week 40, the first week of October, and ends in MMWR Week 39, the last week of September of the following year. **Alt text**: Bar chart showing the distribution of influenza virus subtype and lineages by age groups.

The 2025-26 influenza season was characterized by two distinct waves of activity. The first was driven by influenza A viruses, which accounted for the highest number of positive specimens during the season overall (**Supplementary Figure 3)**. Of influenza A viruses with a subtype determined, 81.9% were influenza A(H3N2). The second wave of virus activity was driven by influenza B viruses. All influenza B viruses with lineage determined belonged to the B/Victoria lineage. These trends were similar across all HHS regions (**Supplementary Figure 4**).

**Figure 3.**
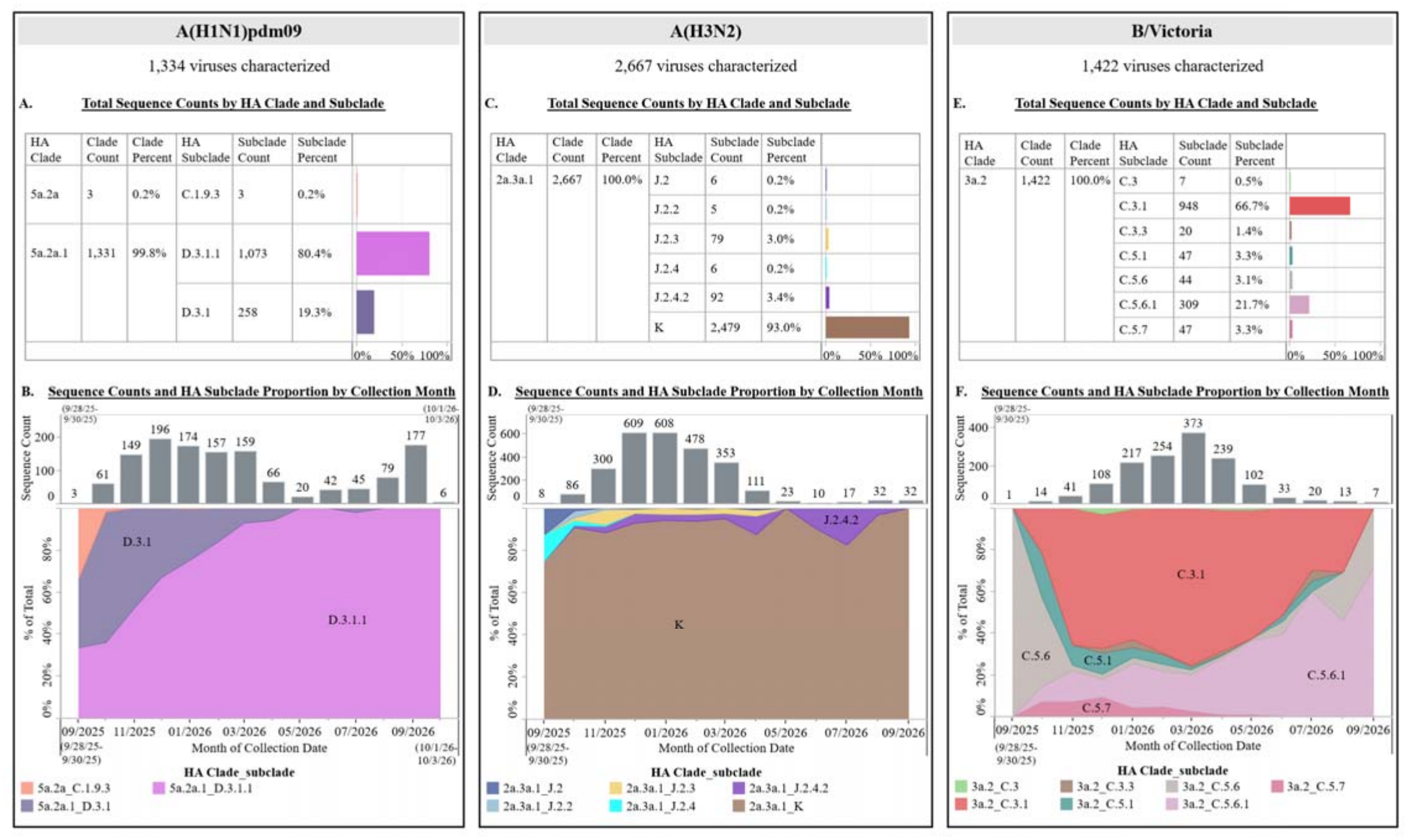
Genetic characterization of hemagglutinin (HA) genes from influenza viruses during the 2025-26 influenza season, United States, September 28, 2025 – October 3, 2026 (Morbidity and Mortality Weekly Report (MMWR)^a^ Weeks 40-39) *A, C, E showed the cumulative sequence counts and percentages by HA clade and subclade for each subtype from influenza viruses collected during the 2025-26 influenza season **B, D, F showed the sequence counts and HA subclade proportion by collection month for each subtype. ^a^MMWR weeks are defined by the Saturday week-ending date. For influenza surveillance, each season begins in MMWR Week 40, the first week of October, and ends in MMWR Week 39, the last week of September of the following year. **Alt text**: Paneled figure with 6 charts (2 per influenza virus type) showing the total sequence counts by hemagglutinin (HA) clade and subclade by influenza viruses A(H1N1), A(H3N2), and B/Victoria and with a graph showing sequence counts and proportions of subclade of each virus by specimen collection month.

**Figure 4.**
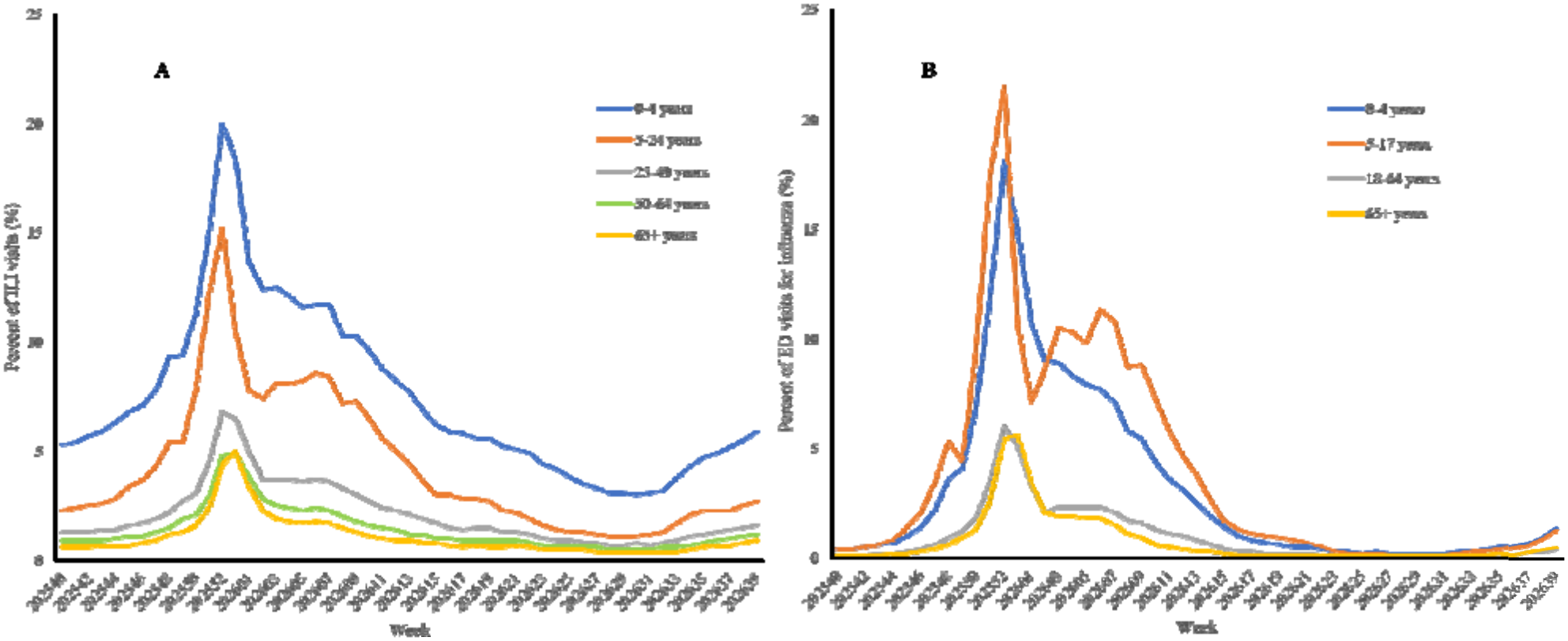
National summary by Morbidity and Mortality Weekly Report (MMWR)^a^ Week of **A**) the percent of outpatient visits for influenza-like illness (ILI) as reported to the U.S. Outpatient Influenza-Like Illness Surveillance Network (ILINet) by age group and **B)** the percent of visits to an emergency department (ED) with a discharge diagnosis of influenza as reported to NSSP by age group during the 2025-26 influenza season, United States, September 28, 2025-October 3, 2026 (MMWR Weeks 40-30) ^a^MMWR weeks are defined by the Saturday week-ending date. For influenza surveillance, each season begins in MMWR Week 40, the first week of October, and ends in MMWR Week 39, the last week of September of the following year. **Alt text**: Paneled line graphs comparing a) the proportion of ILI visits by age group and b) the proportion of ED visits with a discharge diagnosis of influenza by age group by MMWR Week

Of influenza A viruses with subtype information available, influenza A(H3N2) viruses accounted for the largest proportion of influenza viruses detected in each age group, with the highest proportion among persons aged 65 years and older (66.5%), followed by persons aged 0-4 years (59.2%), 25-64 years (52.5%), and 5-24 years (51.7%) (**Figure 2**). In contrast, the proportion of influenza B viruses was highest among persons aged 5-24 years (27.7%), followed by persons aged 25-64 years (17.7%), 0-4 years (13.7%), and 65 years and older (2.6%).

### Novel Influenza A Viruses

Novel influenza A viruses are influenza A virus subtypes different from circulating seasonal influenza A(H1) and A(H3) viruses.^10^ During the 2025-26 season, five confirmed cases of novel influenza A virus infections were reported to CDC. The first known human A(H5N5) virus infection worldwide, was reported by Washington^11,12^ and four variant influenza A(H1N2)v (i.e., a swine influenza virus identified in a human and designated with a “v”) virus infections were reported from Michigan,^13^ Nebraska,^14^ and Vermont^12^. No human-to-human transmission occurred. The influenza A(H5N5) infection was detected through enhanced public health surveillance at the treating hospital. Two influenza A (H1N2)v infections from Michigan were detected through enhanced surveillance associated with an agricultural fair; two of the influenza A(H1N2)v (Vermont and Nebraska) infections were detected through routine virologic surveillance conducted by public health laboratories.

### Virus Characterization and Antiviral Susceptibility

#### Virus Characterization

Following an existing surveillance framework^15^, public health laboratories submitted a subset of influenza-positive specimens to CDC national influenza reference centers for genomic sequencing and virus isolation in cell culture. Hemagglutinin (HA) sequences were classified into clade/subclade and further analyzed to inform the selection of viruses for antigenic characterization at CDC and contribute to a global picture of influenza virus circulation considered during semiannual influenza vaccine strain selection.^16,17^

Cumulatively for the 2025-26 influenza season, CDC genetically characterized 5,423 influenza viruses collected in the U.S.: 1,334 A(H1N1)pdm09 viruses, 2,667 A(H3N2) viruses, and 1,422 B/Victoria lineage viruses (**Figure 3**).

#### Influenza A(H1N1)pdm09

Most HA genes characterized from circulating A(H1N1)pdm09 viruses were HA subclade D.3.1 (19.3%) or D.3.1.1 (80.4%) (**Figure 3**). While D.3.1 was the predominant HA subclade during the 2024-25 influenza season,^18^ D.3.1.1 evolved from D.3.1 and predominated this season. Of 441 A(H1N1)pdm09 viruses antigenically characterized, 432 (98.0%) were antigenically similar to the 2025-26 northern hemisphere A(H1N1)pdm09 vaccine reference virus.^4^

#### Influenza A(H3N2)

All A(H3N2) HA genes of viruses characterized belonged to subclades of J.2 which predominated in the 2024-25 season, but each subclade had acquired several changes in their HA proteins. Viruses from HA subclades J.2, J.2.2, J.2.3, J.2.4 and J.2.4.2 circulated at low levels (each below 4%) (**Figure 3**), while viruses from HA subclade K were predominant (93.0%) throughout this season (**Supplementary Table 1**).^18^ Of 595 A(H3N2) viruses antigenically characterized, only 6 (1.0%) were antigenically similar to the 2025-26 northern hemisphere A(H3N2) vaccine reference virus and the majority belonged to subclades J.2 or J.2.2. The majority of A(H3N2) viruses with subclades J.2.3, J.2.4, J.2.4.2 and K were antigenically drifted from the 2025-26 northern hemisphere A(H3N2) vaccine reference virus.^4^

#### Influenza B/Victoria

Influenza B/Victoria viruses characterized had HA genes from subclades C.3 (C.3, C.3.1, C.3.3) and C.5 (C.5.1, C.5.6, C.5.6.1, C.5.7) (**Figure 3**). Viruses with these subclades also co-circulated during the 2024-25 season.^18^ Subclade C.3.1 HA became the predominant subclade in November 2025 but C.5.6.1 increased in proportion after March 2026. Of 361 B/Victoria viruses antigenically characterized, 131 (36.3 %) were antigenically similar to the 2025-26 northern hemisphere B/Victoria vaccine reference virus.^4^ B/Victoria viruses with C.5 (C.5.1, C.5.6, C.5.6.1, C.5.7) HA were antigenically similar to the 2025-26 northern hemisphere vaccine B/Victoria component, but viruses from HA subclades C.3 (C.3, C.3.1, C.3.3) were antigenically drifted.

#### Antiviral Susceptibility

Based on a combination of sequence-based assessment and in vitro laboratory testing, almost all seasonal influenza viruses collected and assessed for antiviral susceptibility were susceptible to neuraminidase inhibitors oseltamivir, peramivir, and zanamivir and the polymerase acidic inhibitor baloxavir.^19^ Of 1,351 A(H1N1)pdm09 viruses assessed, 10 (0.7%) had an amino acid substitution that conferred highly reduced inhibition by oseltamivir and peramivir; eight were collected from patients without known exposure to oseltamivir or peramivir. Another 28 (2.1%) A(H1N1)pdm09 viruses had amino acid substitutions that conferred reduced inhibition by oseltamivir. Of 2,666 A(H3N2) viruses assessed, two (<0.1%) had an amino acid substitution that conferred highly reduced inhibition by oseltamivir; both were collected from patients treated with oseltamivir. Three (0.2%) of the 1,421 influenza B viruses assessed had an amino acid substitution that conferred reduced inhibition by peramivir and one (<0.1%) had a substitution that conferred reduced inhibition by zanamivir. Only one A(H3N2) virus was identified as having a substitution that conferred reduced susceptibility to baloxavir (**Supplementary Table 2**).

### Outpatient and Emergency Department Visits

Outpatient and emergency department (ED) visits were monitored using data from two surveillance systems: the U.S. Outpatient Influenza-like Illness Surveillance Network (ILINet) and the National Syndromic Surveillance Program (NSSP). ILINet consists of reports from a variety of outpatient healthcare providers, including emergency departments, for influenza-like illness ([ILI], fever plus cough or sore throat) visits, while the data extracted from NSSP for this analysis included ED visits with an influenza discharge diagnosis.^4^

The percent of ILI visits peaked at 8.3% and ED visits with a discharge diagnosis of influenza peaked at 8.6% during the week ending December 27, 2025 (MMWR Week 52). This was five weeks earlier than the 2024-25 season in ILINet and six weeks earlier in NSSP. ILINet remained above the national baseline^3^ of 3.1% for 15 consecutive weeks, and the difference between the ILINet peak and seasonal baseline was the second largest since the 2010-11 season.^5^ Peak timing was consistent across both systems in most HHS regions: HHS regions 1-8 peaked in late December and HHS Region 10 (Pacific Northwest) in early February. Region 9 (West Coast) was the exception, where ILINet peaked in early January and NSSP peaked in early February.

In both systems, the pediatric age groups experienced higher activity than adults. In ILINet, children aged 0-4 years experienced a higher percent of visits for ILI throughout the season compared to persons aged 5-24 years. However, in NSSP children aged 5-17 years experienced a higher peak value and higher values during most weeks of the season for ED visits with a discharge diagnosis of influenza compared to children aged 0-4 years (**Figure 4**).

### Influenza-Associated Hospitalizations

Influenza-associated hospitalizations were monitored through the Influenza Hospitalization Surveillance Network (FluSurv-NET) and the National Healthcare Safety Network (NHSN) Hospital Respiratory Data (HRD) module. FluSurv-NET conducted population-based surveillance on laboratory-confirmed influenza hospitalizations in patients of all ages in more than 90 counties across 14 states and captured individual-level demographic and detailed clinical data.^20^ NHSN reported weekly aggregated numbers of laboratory-confirmed influenza hospitalizations from all acute care and critical access hospitals in the U.S., providing greater geographic coverage but less demographic, laboratory, and clinical data.^21,22^

Both systems showed similar timing and intensity of influenza-associated hospitalizations during the 2025-26 season. Weekly influenza-associated hospitalization rates peaked at 13.1 hospitalizations per 100,000 population in FluSurv-NET during week ending December 27, 2025 (MMWR Week 52); NHSN peaked one week later at 12.6 per 100,000 population. Peaks in both systems were lower and occurred earlier than the 2024-25 season. In both systems, adults aged 65 years and older experienced the highest peak weekly hospitalization rates (45.9 per 100,000 in FluSurv-NET and 44.0 per 100,000 in NHSN), followed by children aged 0-4 years (14.0 and 12.3 per 100,000 population, respectively).

Cumulative hospitalization rates for all ages combined were similar in FluSurv-NET (91.3 per 100,000) and NHSN (101.6 per 100,000). In FluSurv-NET, this was the third-highest cumulative rate observed from October – April since the 2010-11 season (**Supplementary Figure 5**).

**Figure 5.**
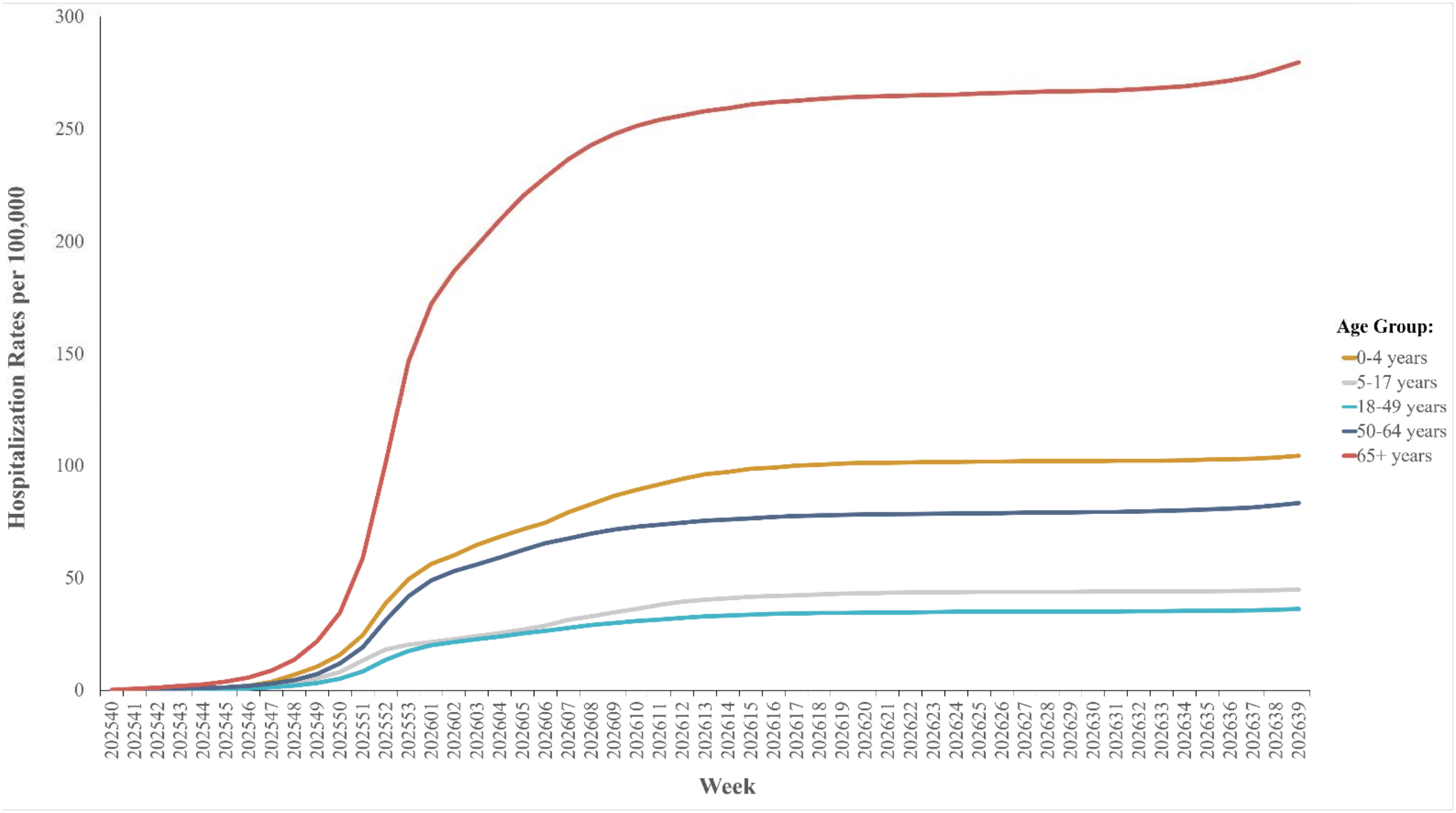
Cumulative rates of laboratory-confirmed influenza-associated hospitalization per 100,000 population by age group as reported to FluSurv-NET during the 2025-26 influenza season, United States, September 28, 2025 – October 3, 2026 (Morbidity and Mortality Weekly Report (MMWR)^a^ Weeks 40-39) ^a^MMWR weeks are defined by the Saturday week-ending date. For influenza surveillance, each season begins in MMWR Week 40, the first week of October, and ends in MMWR Week 39, the last week of September of the following year. **Alt text**: Line graphs comparing the cumulative rates of influenza-associated hospitalizations per 100,000 population by age group as reported to FluSurv-NET by MMWR Week.

Consistent with peak weekly rates, adults aged 65 years and older had the highest cumulative hospitalization rates in both systems (279.2 per 100,000 in FluSurv-NET (**Figure 5**) and 313.6 per 100,000 in NHSN (**Supplementary Figure 6**)), followed by children aged 0-4 years (104.2 and 112.4 per 100,000 population, respectively). The cumulative hospitalization rate for children aged 0-4 years and 5-17 years was the second highest and highest rate for each age group from October – April respectively, since the 2010-11 season; however, for adults the cumulative rate ranged from the third to the sixth highest depending on the age group.

NHSN data showed regional variation in timing of hospitalization peaks. East coast regions and the central U.S (HHS regions 1-8) peaked during late December, while HHS regions 9 and 10 (West Coast and Pacific Northwest, respectively) peaked in late February.

Among all influenza-associated hospitalizations reported in FluSurv-NET, 87.5% were associated with influenza A viruses, 12.2% with influenza B viruses, 0.1% with influenza A virus and influenza B virus coinfection, and 0.2% for which virus type was not determined. Among hospitalizations associated with an influenza A virus and with virus subtype information available, 84.7% of viruses were A(H3N2), and 15.3% were A(H1N1)pdm09.

### Influenza-associated Mortality

Influenza mortality surveillance was conducted through the National Center for Health Statistics (NCHS) Mortality Surveillance System and the Influenza-Associated Pediatric Mortality Surveillance System. Vital statistics offices reported death certificate data to NCHS for nearly all U.S. deaths. Influenza deaths were identified by ICD-10 multiple cause of death codes.^4^ Influenza-associated pediatric mortality is a nationally notifiable condition defined as a death in a child aged 0-17 years, resulting from a clinically compatible illness confirmed to be influenza by an appropriate laboratory diagnostic test; and with no period of complete recovery between illness and death.^23,24^

Of 3,040,798 all-cause deaths reported to NCHS during the 2025-26 season, 10,902 (0.4%) listed influenza as an underlying or contributing cause of death. Influenza deaths increased sharply beginning late December, peaked at 2.0% among all deaths during week ending January 10, 2026 (MMWR Week 1), then declined steadily. This was the third highest peak, following the 2024-25 season (2.8%) and the 2017-18 season (2.5%), since NCHS influenza mortality data were incorporated into influenza surveillance during the 2015-16 influenza season (**Supplementary Figure 7**).

HHS Region 1 had the highest peak percentage of deaths due to influenza at 4.1%, while regions 6 and 9 had the lowest (1.1% and 1.0% respectively). Peaks in regions 1 and 2 were similar (<0.01 percentage point difference) to the 2024-25 season; while all other regions (3-10) reported higher peaks during the 2024-25 season. By age group, children aged 0–18 years had the highest peak (4.2%) during week ending January 3, 2026 (MMWR Week 53), followed by adults aged 65 years and older (2.1%) during week ending January 10, 2026 (MMWR Week 1), and adults aged18–64 years (1.6%) during MMWR Week 53.

During the 2025-26 season, 198 laboratory-confirmed influenza-associated pediatric deaths were reported to CDC marking the fourth highest seasonal total since reporting began in 2004, excluding the 2009-10 influenza A(H1N1)pdm09 pandemic (**Supplementary Figure 8**). Of these deaths, 137 (69.2%) were associated with influenza A viruses and 61 (30.8%) were associated with influenza B viruses. Among 95 influenza A virus infections with subtype reported, 81 (85.3%) were A(H3N2) and 14 (14.7%) were A(H1N1).

### Influenza Disease Burden and Severity

#### Disease Burden and Severity

Influenza surveillance systems are not designed to identify all cases of influenza virus infection.^25^ Therefore, age-specific hospitalization rates from FluSurv-NET were used in a mathematical model to estimate the number of influenza-associated illnesses, medical visits, hospitalizations, and deaths, adjusted for under detection of influenza in FluSurv-NET.^26,27^ A Moving Epidemic Model utilized surveillance data for influenza-like illnesses, hospitalizations, and deaths for all age groups to classify seasonal severity.^28,29^

During the 2025-26 season, CDC estimated there were between 30 million – 49 million illnesses, 14 million – 23 million medical visits, 360,000 – 690,000 hospitalizations, and 20,000 – 48,000 deaths related to influenza (**Supplementary Table 3a**). CDC classified the overall 2025-26 season severity as moderate.^30^ When stratified by age group, children experienced a high-severity season, while adults experienced a moderate-severity season.

#### Prevented Influenza Disease Burden from Vaccination

The total number of influenza illnesses, medical visits, hospitalizations, and deaths prevented by influenza vaccination were modelled by incorporating season-specific data on burden of disease, influenza vaccine coverage, and vaccine effectiveness (VE).^31,32^ It was estimated between 3.8 million – 6.8 million illnesses, 1.9 million – 3.4 million medical visits, 60,000 – 130,000 hospitalizations, and 3,600 – 10,000 deaths were prevented by influenza vaccination during the 2025-26 season (**Supplementary Table 3b**).

## Discussion

Influenza A(H3N2) viruses were predominant for the 2025-26 season overall. More than 90% of genetically characterized A(H3N2) viruses belonged to the HA subclade K and were antigenically drifted from the corresponding 2025-26 influenza vaccine reference viruses.^33^ National influenza activity peaked in late December 2025, five weeks earlier than the 2024-25 season peak, but followed a seasonal pattern within historic norms.^34,18^ Influenza activity peaked first in the eastern and central U.S., with HHS regions 1-8 peaking in late December, while western regions (HHS regions 9 and 10) experienced later peaks in early February. Despite these regional differences, the distribution of influenza types and subtypes were similar across the nation.

Influenza B viruses increased during the latter part of the season, producing a second wave of activity consistent with some previous influenza seasons.^5^ Slightly over half of antigenically characterized influenza B/Victoria viruses were antigenically drifted from the corresponding 2025–26 influenza vaccine reference viruses. The circulation of antigenically drifted influenza viruses highlights the importance of virologic surveillance to monitor changes in circulating influenza viruses and inform vaccine strain selection recommendations, as was evident in the recommendation to update all three components of the 2026–27 Northern Hemisphere influenza vaccine.^35^ Despite antigenic changes, circulating influenza viruses remained largely susceptible to recommended antiviral medications and influenza vaccination was estimated to have prevented millions of influenza-associated illnesses.^36,37^ Additionally, while only five cases of novel influenza A virus infection were identified, these detections reinforce the importance of maintaining comprehensive year-round influenza surveillance including subtyping of influenza A viruses and thorough investigation of novel virus infections to ensure person-to-person transmission is not occurring.

Like prior influenza seasons, children experienced higher outpatient visits for ILI and ED visits with a discharge diagnosis of influenza, and peak percentage of influenza coded deaths compared to the adult age groups, while older adults experienced the highest rate of influenza-associated hospitalizations compared to other age groups. The percentage of visits for ILI was higher most weeks among children 0-4 years than those 5-24 years while the percent of ED visits with a discharge diagnosis of influenza was higher most weeks in those 5-17 years compared to those 0-4 years. While both systems track generally less severe medically attended illness, the difference between which age groups had higher activity may be due to ILINet having a broader age group for children which includes persons aged 5-24 years, while NSSP has a smaller pediatric age group including children aged 5-17 years. Additionally, ILINet tracks visits for syndromes that could also be caused by other respiratory viruses such as respiratory syncytial virus (RSV), a respiratory virus known to disproportionately affect younger children.^38^

Although peak weekly influenza-associated hospitalization rates were above or similar to historic norms for all age groups, evaluating cumulative rates and estimates of influenza burden provides a more complete assessment of the impact of influenza, as peak activity can be impacted by the timing of activity across the country and does not fully capture the full impact of influenza over the course of a season. Adults 65 years and older experienced the highest cumulative hospitalization rate compared to other age groups, but these rates were within historic norms for all adult age groups.^39^ For children aged 0-4 years and 5-17 years, the cumulative hospitalization rate was second highest and highest, respectively for these age groups, compared to prior seasons since the 2010-11 season.

Influenza A(H3N2) HA subclade K viruses predominated during the 2025-26 influenza season and a wave of influenza B occurred later in the season. The season severity was classified as moderate overall and for adults, but high for children. Continued integration of virologic, epidemiologic, and clinical surveillance remains essential for detecting novel influenza A viruses, monitoring changes in seasonal influenza virus circulation, informing seasonal influenza vaccine composition, and guiding public health interventions.

## Acknowledgments

We would like to thank Amara Fazal, Angelle Kardorff, Camden Castagna-McLeod, and Charisse Cummings. We would also like to thank state, county, city, and territorial health departments and public health laboratories; CDC Influenza Division Staff; Influenza collaborating laboratories; National Respiratory and Enteric Virus Surveillance System laboratories; The Association of Public Health Laboratories; National Influenza Reference Centers; U.S. Outpatient Influenza-Like Illness Surveillance (ILI) Network sites; the National Syndromic Surveillance Program (NSSP); Influenza Hospitalization Surveillance Network (FluSurv-NET) sites; the National Healthcare Safety Network (NHSN); Division of Healthcare Quality Promotion, CDC; National Center for Health Statistics, CDC; National Center for Immunization and Respiratory Diseases, CDC.

## Data availability statement

The data reported in *FluView* including ILINet, clinical and public health laboratories, FluSurv-NET and NHSN hospitalizations, novel influenza A, and mortality data are accessible through <u>FluView Interactive | FluView | CDC</u>.

NSSP emergency department data are accessible through <u>NSSP Emergency Department Visit Trajectories by State and Sub State Regions-COVID-19, Flu, RSV, Combined□ □ | Data | Centers for Disease Control and Prevention</u>

