## Supplemental materials for "Influenza activity in the United States during the 2025-26 influenza season"

**Supplementary Table 1.** Amino acid substitutions compared to the J.2 subclade base consensus hemagglutinin (HA) for the base consensus HA belonging to each HA subclade circulating during the 2025-26 influenza season, United States, September 28, 2025- October 3, 2026 (Morbidity and Mortality Weekly Report (MMWR)^a^ Weeks 40-39)

| **Subclade** | **Amino Acid Substitutions of the Subclade Base Consensus HA Compared to the J.2 Subclade Base Consensus HA** |
| --- | --- |
| **J.2.2** | S124N |
| **J.2.3** | N158K, K189R, S378N |
| **J.2.4** | T135K, K189R |
| **J.2.4.2** | F79V, T135K, S144N, N158D, I160K, K189R, T328A |
| **K** | K2N, T135K, S144N, N158D, I160K, Q173R, K189R, T328A, S378N |

^a^MMWR weeks are defined by the Saturday week-ending date. For influenza surveillance, each season begins in MMWR Week 40, the first week of October, and ends in MMWR Week 39, the last week of September of the following year

**Supplementary Table 2**. Antiviral susceptibility of influenza viruses to neuraminidase (NA) inhibitors and polymerase acidic (PA) cap-dependent endonuclease inhibitor during the 2025-26 influenza season, United States, September 28, 2025- October 3, 2026 (Morbidity and Mortality Weekly Report (MMWR)^a^ Weeks 40-39)

| **Antiviral Medication** | | | **Total Viruses** | **A(H1)** | **A(H3)** | **B/Victoria** |
| --- | --- | --- | --- | --- | --- | --- |
| **Neuraminidase Inhibitors** | **Oseltamivir** | **Viruses Tested** | 5438 | 1351 | 2666 | 1421 |
|  |  | **Reduced Inhibition** | 28 (0.5%) | 28 (2.1%) | 0 | 0 |
|  |  | **Highly Reduced Inhibition** | 12 (0.2%) | 10 (0.7%) | 2 (<0.1%) | 0 |
|  | **Peramivir** | **Viruses Tested** | 5438 | 1351 | 2666 | 1421 |
|  |  | **Reduced Inhibition** | 3 (<0.1%) | 0 | 0 | 3 (0.2%) |
|  |  | **Highly Reduced Inhibition** | 10 (0.2%) | 10 (0.7%) | 0 | 0 |
|  | **Zanamivir** | **Viruses Tested** | 5438 | 1351 | 2666 | 1421 |
|  |  | **Reduced Inhibition** | 1 (<0.1%) | 0 | 0 | 1 (<0.1%) |
|  |  | **Highly Reduced Inhibition** | 0 | 0 | 0 | 0 |
| **PA Cap-Dependent Endonuclease Inhibitor** | **Baloxavir** | **Viruses Tested** | 5282 | 1286 | 2596 | 1400 |
|  |  | **Decreased Susceptibility** | 1 (<0.1%) | 0 | 1 (<0.1%) | 0 |

Ten A(H1N1)pdm09 viruses had NA-H275Y amino acid substitution conferring highly reduced inhibition by oseltamivir and peramivir. Twenty-eight A(H1N1)pdm09 viruses were detected with a combination of NA substitutions, S247N+I223V (n = 27) or S247N+I223T (n =1), conferring reduced inhibition by oseltamivir. Two A(H3N2) viruses had NA-E119V amino acid substitution associated with highly reduced inhibition by oseltamivir. Four B/Victoria viruses had NA substitution, M464T (n=3) and G407S (n=1), conferring reduced inhibition by peramivir and zanamivir, respectively. Only one A(H3N2) virus had PA-K34R, conferring reduced susceptibility to baloxavir.

^a^MMWR weeks are defined by the Saturday week-ending date. For influenza surveillance, each season begins in MMWR Week 40, the first week of October, and ends in MMWR Week 39, the last week of September of the following year.

**Supplementary Figure 1:** National percent of specimens testing positive for any influenza virus (percent positive) by Morbidity and Mortality Weekly Report (MMWR)^a^ Week as reported to the Centers for Disease Control and Prevention (CDC) by a subset of United States clinical laboratories, 2015-16 through 2025-26 influenza seasons

Percent Positive (%)

| Week | 2015-16 season | 2016-17 season | 2017-18 season | 2018-19 season | 2019-20 season | 2020-21 season | 2021-22 season | 2022-23 season | 2023-24 season | 2024-25 season | 2025-26 season |
| --- | --- | --- | --- | --- | --- | --- | --- | --- | --- | --- | --- |
| 40 | 1.05578 | 1.56951 | 2.20191 | 1.74855 | 1.30976 | 0.327869 | 0.15698 | 2.66968 | 1.10103 | 0.674072 | 0.524646 |
| 41 | 1.29662 | 1.35914 | 2.08975 | 1.6967 | 1.47877 | 0.388636 | 0.120916 | 3.46261 | 1.34933 | 0.671305 | 0.536273 |
| 42 | 1.10855 | 1.40304 | 2.17647 | 1.99298 | 1.55203 | 0.350715 | 0.132889 | 5.43662 | 1.51345 | 0.725634 | 0.644416 |
| 43 | 1.10807 | 1.50862 | 2.58279 | 2.05527 | 2.25335 | 0.252845 | 0.254267 | 8.37742 | 1.81282 | 0.964298 | 0.78546 |
| 44 | 1.12344 | 1.91569 | 3.60729 | 2.17372 | 3.05701 | 0.199494 | 0.282283 | 12.7953 | 2.61941 | 1.21851 | 1.09444 |
| 45 | 1.38224 | 2.20089 | 4.24472 | 2.7331 | 5.16261 | 0.205981 | 0.545299 | 14.9355 | 3.95879 | 1.52773 | 1.90515 |
| 46 | 1.19344 | 2.57608 | 5.29966 | 3.15674 | 6.75594 | 0.217721 | 1.00405 | 18.3419 | 4.86504 | 2.11081 | 3.26408 |
| 47 | 1.38506 | 3.34773 | 7.0877 | 3.92782 | 9.54599 | 0.22712 | 1.84175 | 25.8135 | 5.91987 | 2.83398 | 5.47544 |
| 48 | 1.39498 | 3.3191 | 7.30533 | 3.91485 | 10.9385 | 0.155913 | 2.89274 | 25.5531 | 6.96915 | 3.94831 | 8.02224 |
| 49 | 1.47544 | 4.25987 | 10.7453 | 6.23152 | 11.6554 | 0.225744 | 4.05392 | 26.2658 | 9.79897 | 5.15168 | 8.98526 |
| 50 | 2.51181 | 6.68342 | 15.3549 | 10.3644 | 16.1542 | 0.244148 | 6.23128 | 24.4693 | 13.3872 | 8.59373 | 15.4562 |
| 51 | 2.287 | 10.7819 | 22.777 | 14.2649 | 22.533 | 0.285945 | 5.76656 | 20.7763 | 17.3213 | 14.1794 | 25.5832 |
| 52 | 2.45958 | 13.9993 | 25.3864 | 16.352 | 26.9336 | 0.27816 | 4.2145 | 14.8994 | 18.1605 | 18.2344 | 31.7039 |
| 53 | 2.69547 | 13.6715 | 25.3758 | 14.2453 | 25.211 | 0.264387 | 3.32484 | 11.7558 | 15.9818 | 18.2263 | 24.6931 |
| 01 | 2.93137 | 13.3436 | 25.3653 | 12.1387 | 23.4883 | 0.27351 | 2.43518 | 8.61225 | 13.8031 | 18.2181 | 18.6029 |
| 02 | 4.25384 | 15.373 | 26.9421 | 12.7217 | 23.1187 | 0.17222 | 2.06453 | 4.58566 | 13.867 | 18.3976 | 17.0323 |
| 03 | 5.48463 | 18.2865 | 27.034 | 16.3174 | 26.0826 | 0.21281 | 2.11855 | 2.94035 | 14.9059 | 23.8246 | 17.5167 |
| 04 | 6.95974 | 18.5299 | 27.3698 | 19.3918 | 28.2813 | 0.170073 | 2.02929 | 2.12014 | 16.1303 | 28.7465 | 17.4104 |
| 05 | 9.69858 | 21.4215 | 27.0643 | 22.5493 | 30.1465 | 0.202413 | 2.33841 | 1.67339 | 16.3067 | 31.6481 | 18.0427 |
| 06 | 12.5491 | 24.1525 | 26.9981 | 25.1342 | 30.2596 | 0.134941 | 2.93519 | 1.36967 | 15.8521 | 31.4103 | 18.3248 |
| 07 | 15.5359 | 24.5117 | 26.1174 | 26.026 | 29.675 | 0.12645 | 4.11418 | 1.08468 | 15.5673 | 27.8373 | 18.0222 |
| 08 | 18.3621 | 24.7251 | 22.6155 | 26.2407 | 28.3215 | 0.122349 | 4.6558 | 1.01838 | 15.5181 | 23.9403 | 15.6738 |
| 09 | 21.1098 | 19.772 | 18.4867 | 26.0743 | 25.7517 | 0.0906857 | 5.98405 | 0.92233 | 14.9467 | 19.4796 | 15.3439 |
| 10 | 23.6454 | 19.2714 | 15.6938 | 25.6065 | 22.4914 | 0.105291 | 6.86997 | 1.04196 | 14.2261 | 15.8557 | 13.8166 |
| 11 | 19.972 | 19.0338 | 15.5813 | 26.1318 | 15.8125 | 0.129758 | 7.99444 | 0.89434 | 11.9214 | 13.3136 | 11.2108 |
| 12 | 18.4709 | 19.7113 | 15.328 | 22.4805 | 7.50171 | 0.0838856 | 8.28915 | 0.949267 | 10.3628 | 10.7287 | 9.61931 |
| 13 | 16.2265 | 18.4816 | 15.1135 | 19.3035 | 2.32158 | 0.0689655 | 8.55245 | 0.959683 | 9.15264 | 9.2285 | 7.71154 |
| 14 | 14.0164 | 15.4251 | 12.6888 | 14.9422 | 1.0312 | 0.0761421 | 9.49544 | 1.05613 | 7.53799 | 7.57976 | 5.85926 |
| 15 | 13.2362 | 12.7395 | 11.2486 | 11.9093 | 0.61823 | 0.069449 | 9.88646 | 1.01401 | 5.79303 | 6.16646 | 4.51384 |
| 16 | 12.3464 | 9.69626 | 9.39813 | 8.61102 | 0.623139 | 0.0915991 | 8.83682 | 0.837728 | 4.7576 | 5.2814 | 3.41479 |
| 17 | 10.2615 | 6.76776 | 7.99876 | 5.84355 | 0.218375 | 0.083208 | 8.15632 | 0.954306 | 3.79997 | 4.42023 | 2.96964 |
| 18 | 8.12094 | 5.91752 | 6.25914 | 4.81976 | 0.262953 | 0.0828473 | 8.45266 | 0.943503 | 3.1963 | 3.34042 | 2.47606 |
| 19 | 6.68559 | 5.33264 | 4.39311 | 3.83986 | 0.326173 | 0.103324 | 7.27228 | 1.01804 | 2.3264 | 2.6225 | 1.90845 |
| 20 | 5.81108 | 4.86286 | 3.16606 | 3.54159 | 0.305966 | 0.151323 | 6.00416 | 1.17603 | 1.82353 | 2.14267 | 1.5432 |
| 21 | 4.71918 | 4.35223 | 2.39003 | 3.41968 | 0.212681 | 0.150566 | 5.78813 | 1.19412 | 1.64013 | 1.92089 | 1.30898 |
| 22 | 3.0595 | 4.16524 | 1.52934 | 3.0826 | 0.16518 | 0.0921042 | 4.90161 | 1.30539 | 1.63898 | 1.8643 | 0.913254 |
| 23 | 3.02006 | 3.38586 | 1.57683 | 2.78989 | 0.339751 | 0.102938 | 3.31143 | 1.0945 | 1.47982 | 1.59902 | 0.789391 |
| 24 | 1.82927 | 3.06229 | 1.29914 | 2.31579 | 0.279818 | 0.118658 | 2.36341 | 1.10746 | 1.28263 | 1.20473 | 0.642405 |
| 25 | 1.71228 | 2.64932 | 1.02329 | 1.90194 | 0.38117 | 0.129661 | 1.68997 | 0.947234 | 1.04808 | 1.03725 | 0.47613 |
| 26 | 1.22261 | 2.53401 | 1.11356 | 2.0806 | 0.282336 | 0.0703111 | 1.26806 | 1.10448 | 0.890646 | 0.848557 | 0.554731 |
| 27 | 0.903312 | 2.17791 | 1.00305 | 2.42883 | 0.207734 | 0.055857 | 0.973498 | 1.20836 | 0.986472 | 0.797926 | 0.75958 |
| 28 | 0.869153 | 2.16392 | 0.916118 | 2.01653 | 0.173996 | 0.106646 | 0.792748 | 0.998011 | 0.830267 | 0.780353 | 0.544419 |
| 29 | 0.849185 | 1.83895 | 1.0534 | 2.21849 | 0.372409 | 0.0649398 | 0.62007 | 0.998153 | 0.692818 | 0.541867 | 0.454347 |
| 30 | 0.781793 | 1.80607 | 0.995099 | 2.37706 | 0.148957 | 0.0759678 | 0.516078 | 0.994042 | 0.602587 | 0.408367 | 0.429637 |
| 31 | 0.933921 | 1.94796 | 0.953592 | 2.39817 | 0.13104 | 0.110769 | 0.490616 | 1.06399 | 0.558449 | 0.4421 | 0.485751 |
| 32 | 0.900745 | 1.90048 | 0.95729 | 2.05446 | 0.173776 | 0.0539911 | 0.481129 | 0.855174 | 0.485156 | 0.3833 | 0.669173 |
| 33 | 0.803482 | 1.34281 | 0.764331 | 2.08183 | 0.130719 | 0.071706 | 0.479156 | 0.897271 | 0.451414 | 0.34551 | 0.794947 |
| 34 | 1.40485 | 1.43382 | 1.33625 | 2.36167 | 0.224431 | 0.0598399 | 0.593085 | 0.769782 | 0.436722 | 0.388672 | 1.01232 |
| 35 | 1.67771 | 1.93535 | 1.50367 | 3.45517 | 0.310414 | 0.0851229 | 0.616677 | 0.718069 | 0.415591 | 0.397262 | 1.46109 |
| 36 | 1.46146 | 1.88806 | 1.74739 | 3.09749 | 0.198686 | 0.0541829 | 0.753163 | 0.701222 | 0.440743 | 0.391413 | 1.71249 |
| 37 | 1.51255 | 1.89622 | 1.68745 | 2.48391 | 0.184162 | 0.0574581 | 0.867205 | 0.792616 | 0.476351 | 0.451066 | 2.52224 |
| 38 | 1.74135 | 1.66942 | 1.69929 | 2.75656 | 0.395893 | 0.0713688 | 1.24527 | 0.915555 | 0.531351 | 0.463353 | 3.05455 |
| 39 | 1.78369 | 1.70313 | 1.49699 | 2.74367 | 0.221754 | 0.117498 | 2.01952 | 0.933506 | 0.529423 | 0.642488 | 3.85891 |

^a^MMWR weeks are defined by the Saturday week-ending date. For influenza surveillance, each season begins in MMWR Week 40, the first week of October, and ends in MMWR Week 39, the last week of September of the following year

**Supplementary Figure 2.** Regional distribution (HHS Regions 1-10) ^a^ of the number of positive influenza A and B specimens and percent positivity for all influenza viruses, influenza A, and influenza B as reported to the Centers for Disease Control and Prevention (CDC) by a subset of clinical laboratories in the United States, September 28, 2025- October 3, 2026 (Morbidity and Mortality Weekly Report (MMWR)^b^ Weeks 40-39)

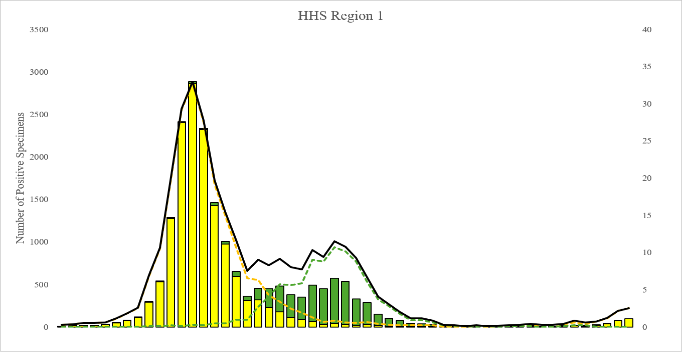

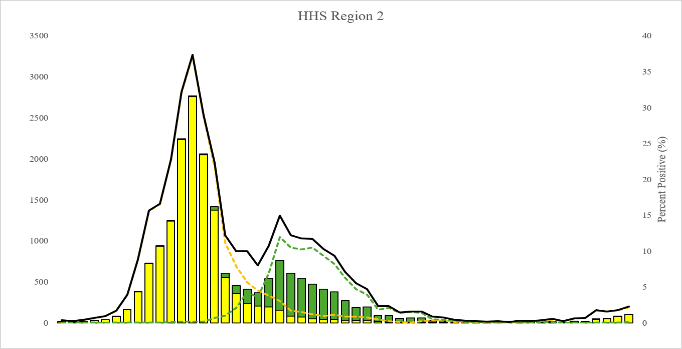

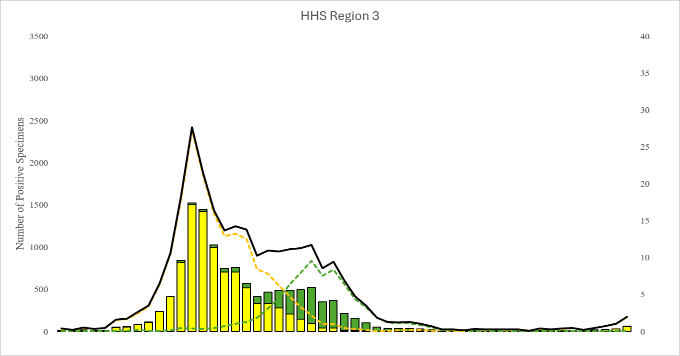

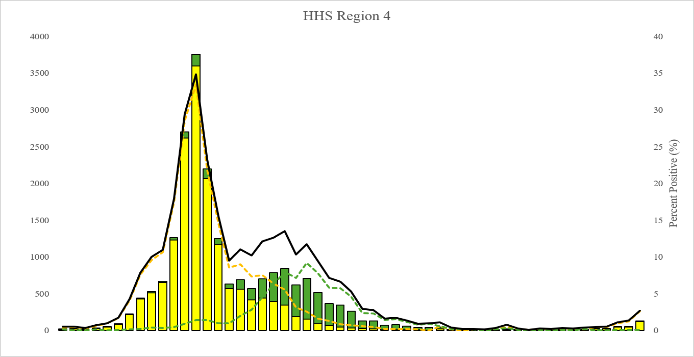

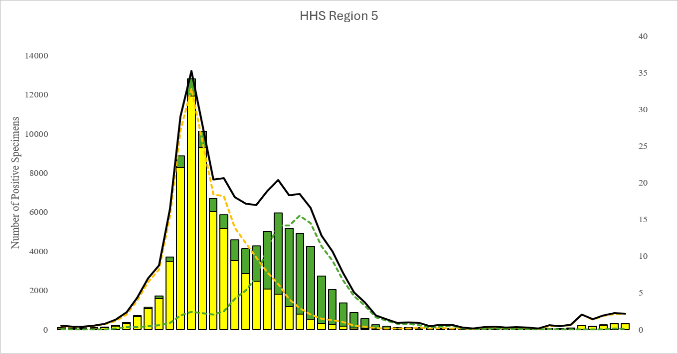

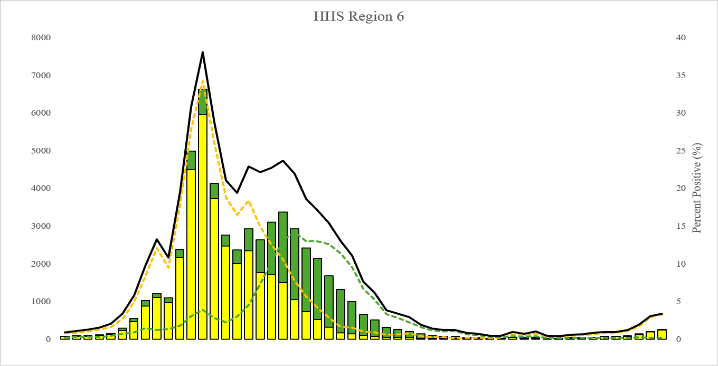

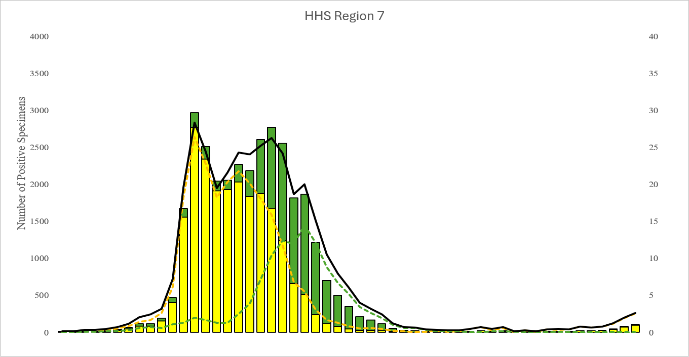

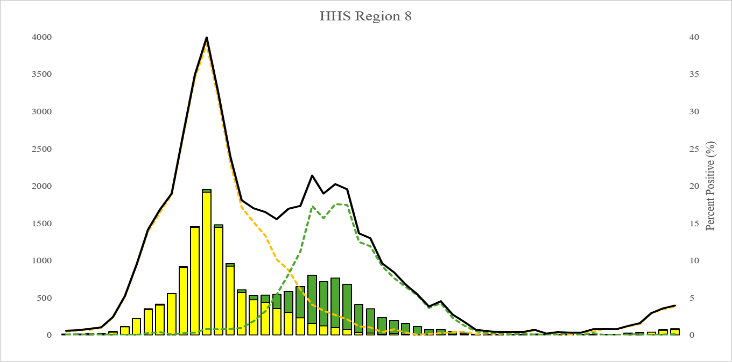

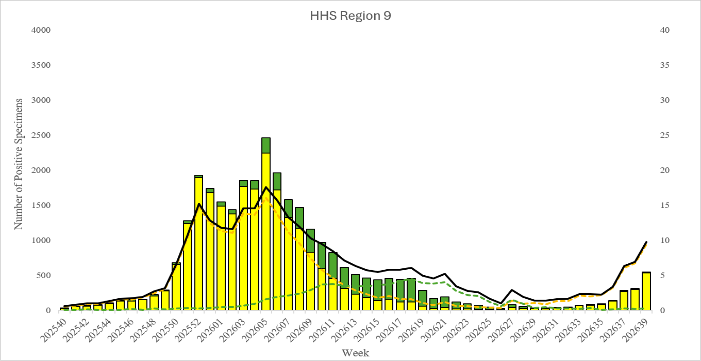

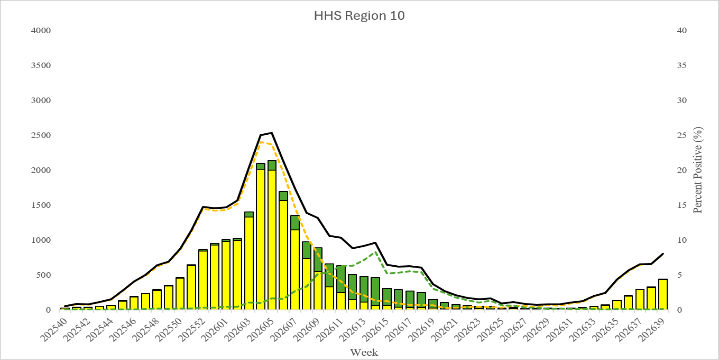

^a^HHS regions are defined as: Region 1 (New England), Region 2 (New York/New Jersey/Puerto Rico), Region 3 (Mid-Atlantic), Region 4 (Southeast), Region 5 (Midwest), Region 6 (South Central), Region 7 (Central), Region 8 (Mountain), Region 9 (West Coast), and Region 10 (Pacific Northwest)

^b^MMWR weeks are defined by the Saturday week-ending date. For influenza surveillance, each season begins in MMWR Week 40, the first week of October, and ends in MMWR Week 39, the last week of September of the following year.

**Supplementary Figure 3**. National distribution of influenza viruses by Morbidity and Mortality Weekly Report (MMWR)^a^ Week as reported the Centers for Disease Control and Prevention (CDC) by public health laboratories, United States, September 28, 2025- October 3, 2026 (MMWR Weeks 40-39)

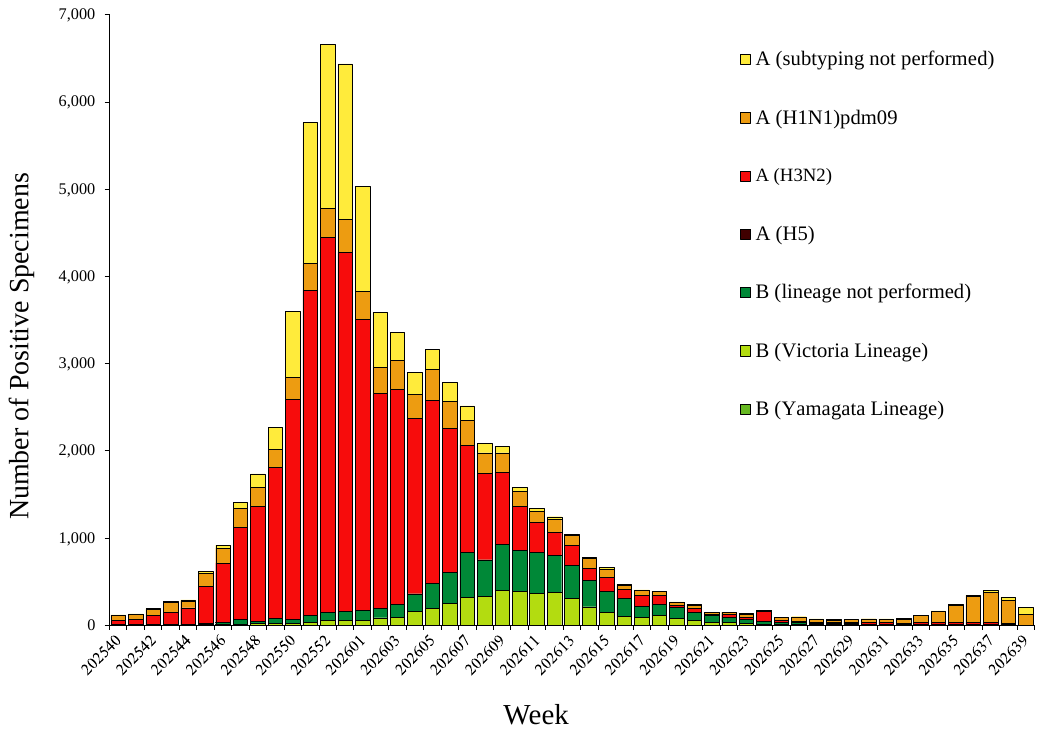

^a^MMWR weeks are defined by the Saturday week-ending date. For influenza surveillance, each season begins in MMWR Week 40, the first week of October, and ends in MMWR Week 39, the last week of September of the following year.

**Supplementary Figure 4.** Regional (HHS Regions 1-10)^a^ proportions of influenza viruses as reported to the Centers for Disease Control and Prevention (CDC), by public health laboratories, United States, September 28, 2025- October 3, 2026 (Morbidity and Mortality Weekly Report (MMWR)^b^ Weeks 40-39)

Proportion of Viruses (%)

| HHS Regions | A (H3N2) (%) | A (H1N1pdm09) (%) | A Unknown (%) | Influenza B (%) |
| --- | --- | --- | --- | --- |
| 1 | 61.8657 | 8.75349 | 13.5983 | 15.7826 |
| 2 | 78.3409 | 11.3157 | 1.51014 | 8.83326 |
| 3 | 38.4348 | 4.96084 | 46.1338 | 10.4706 |
| 4 | 64.8672 | 15.865 | 2.21106 | 17.0567 |
| 5 | 57.8816 | 10.0833 | 13.0585 | 18.9765 |
| 6 | 63.5423 | 11.4134 | 4.87877 | 20.1656 |
| 7 | 72.0571 | 8.56812 | 1.42802 | 17.9467 |
| 8 | 59.6444 | 11.0961 | 1.14787 | 28.1116 |
| 9 | 55.8975 | 22.1881 | 4.97678 | 16.9376 |
| 10 | 62.1394 | 21.4657 | 1.93514 | 14.4598 |

^a^HHS regions are defined as: Region 1 (New England), Region 2 (New York/New Jersey/Puerto Rico), Region 3 (Mid-Atlantic), Region 4 (Southeast), Region 5 (Midwest), Region 6 (South Central), Region 7 (Central), Region 8 (Mountain), Region 9 (West Coast), and Region 10 (Pacific Northwest)

^b^MMWR weeks are defined by the Saturday week-ending date. For influenza surveillance, each season begins in MMWR Week 40, the first week of October, and ends in MMWR Week 39, the last week of September of the following year.

**Supplementary Figure 5**. Cumulative rates of laboratory-confirmed influenza-associated hospitalizations per 100,000 population as reported to FluSurv-NET, 2010-11 through 2025-26 influenza seasons, United States, September 28, 2025- October 3, 2026 (Morbidity and Mortality Weekly Report (MMWR)^a^ Weeks 40-39)

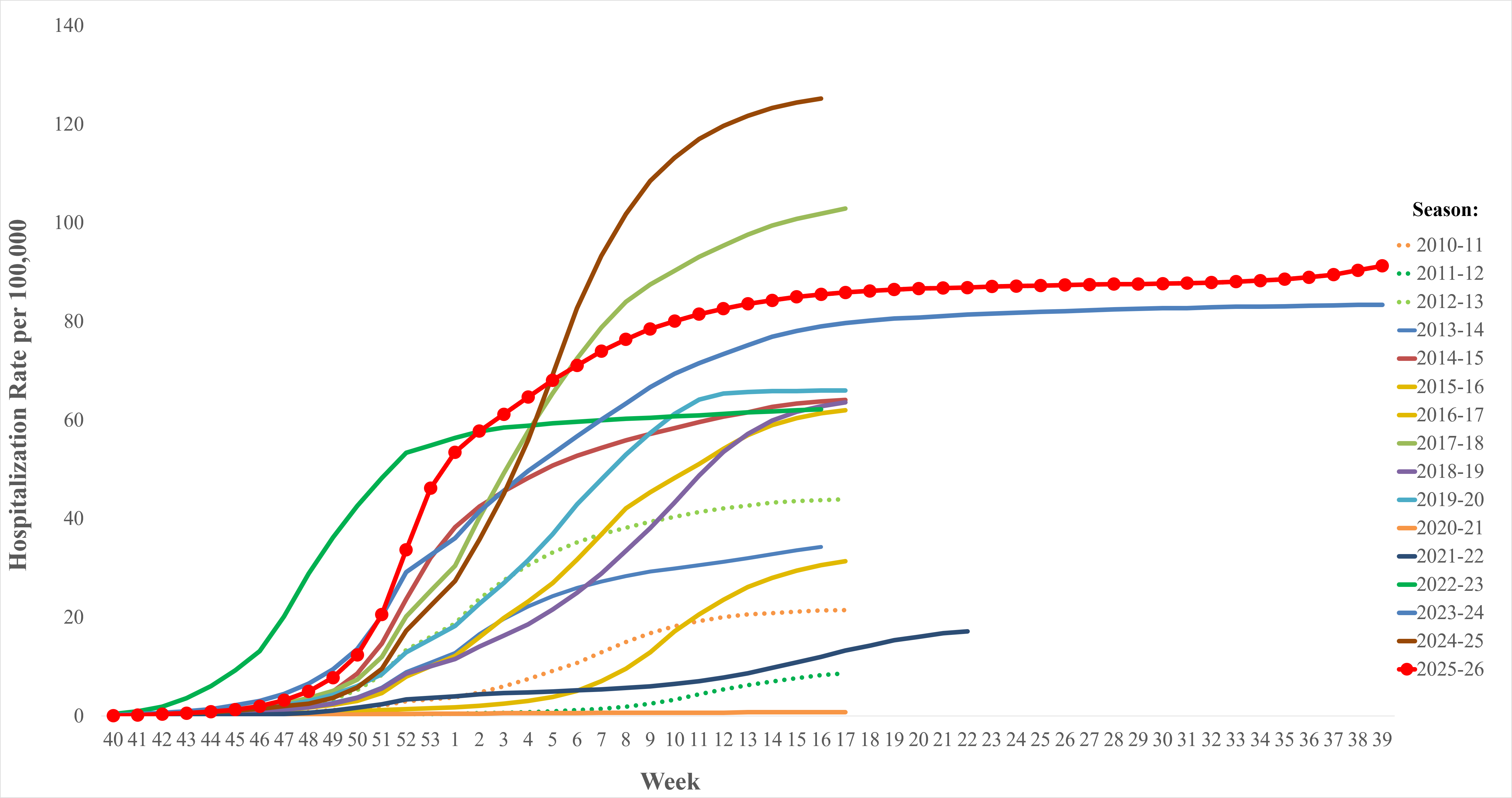

^a^MMWR weeks are defined by the Saturday week-ending date. For influenza surveillance, each season begins in MMWR Week 40, the first week of October, and ends in MMWR Week 39, the last week of September of the following year

**Supplementary Figure 6**: Weekly influenza-associated hospitalization rates by age group reported to National Health Safety Network by Morbidity and Mortality Weekly Report (MMWR)^a^ Week during the 2025-26 influenza season, United States, September 28, 2025- October 3, 2026 (MMWR Weeks 40-39)

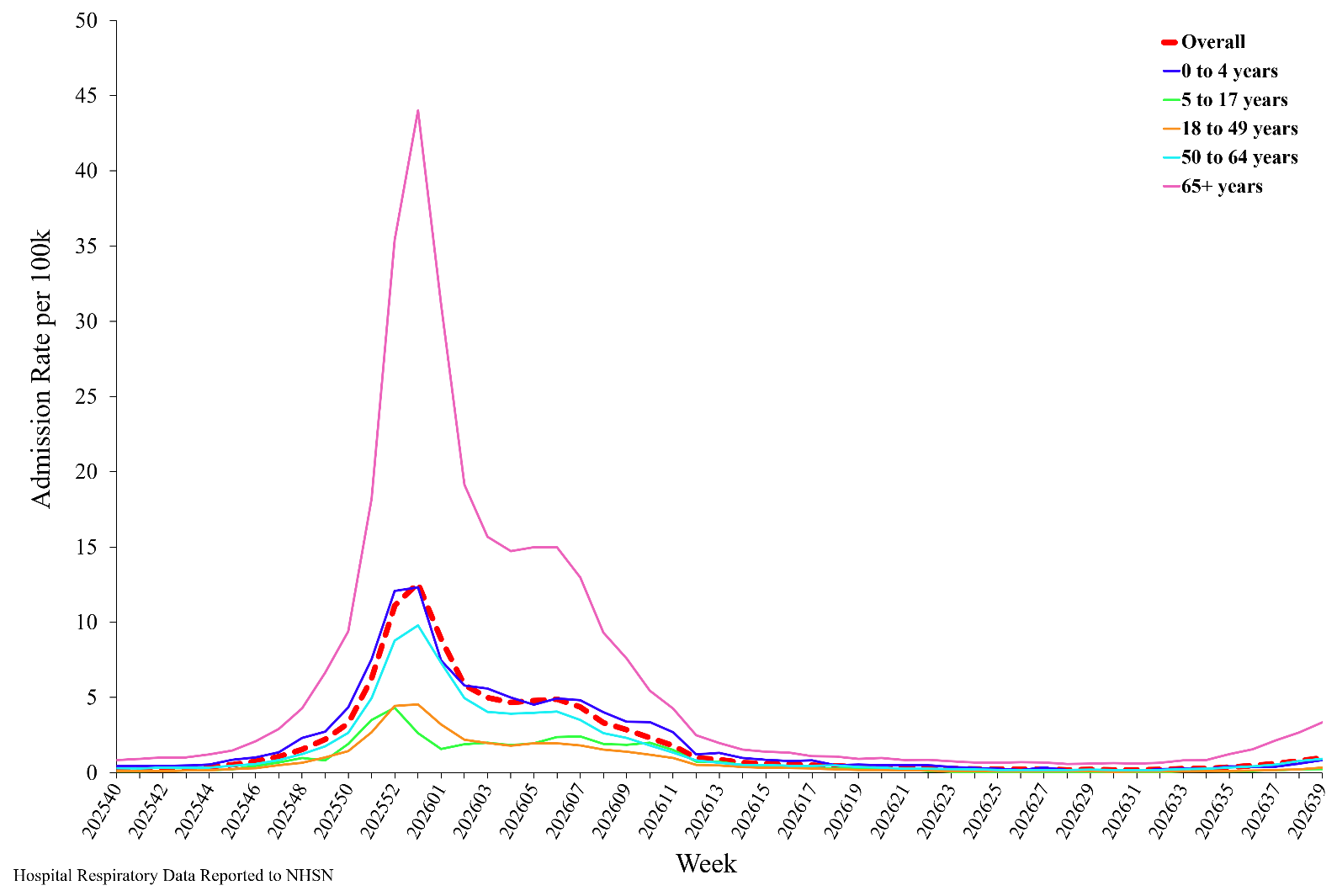

^a^MMWR weeks are defined by the Saturday week-ending date. For influenza surveillance, each season begins in MMWR Week 40, the first week of October, and ends in MMWR Week 39, the last week of September of the following year

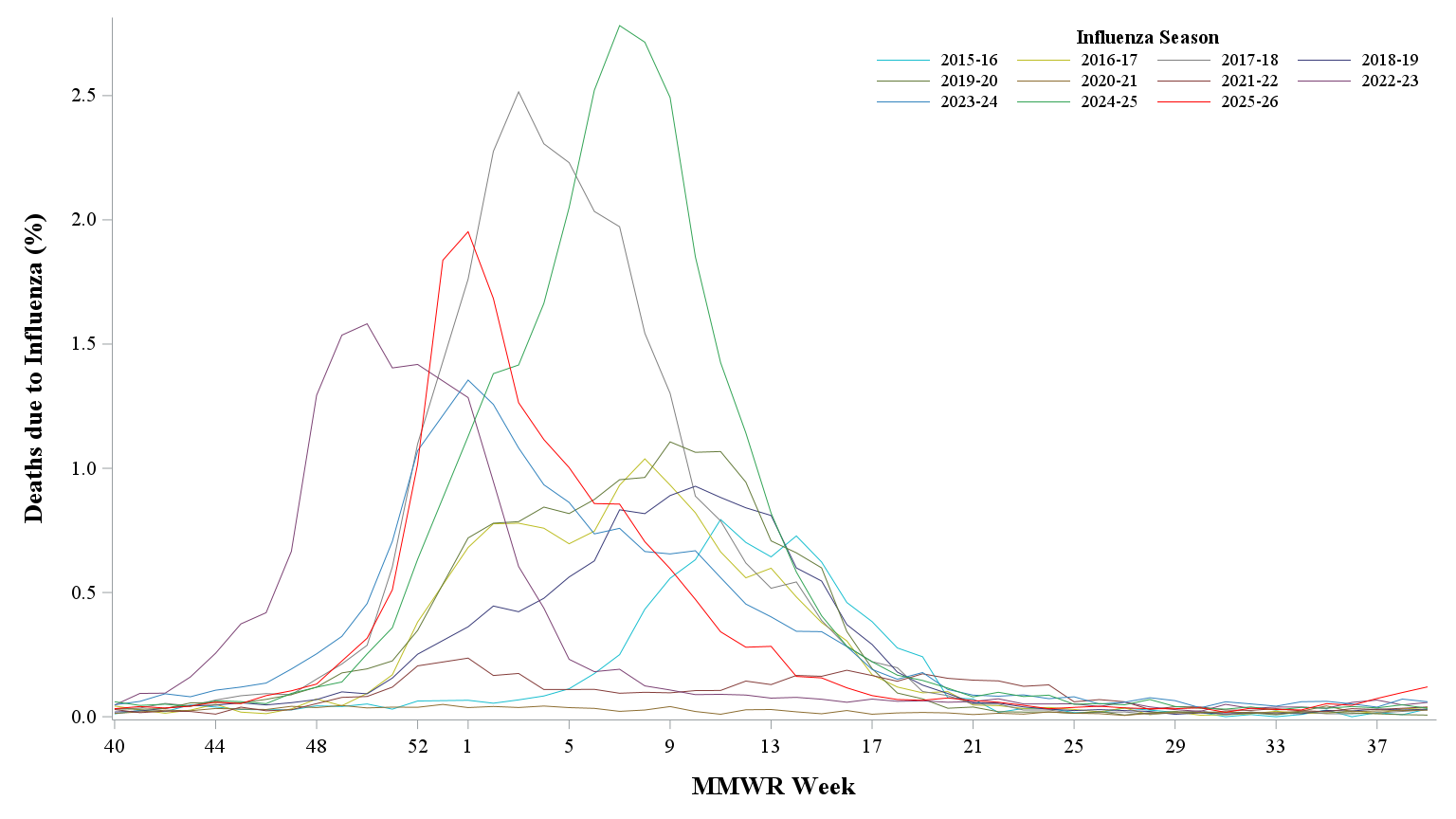
**Supplementary Figure 7**: National summary of influenza coded deaths by Morbidity and Mortality Weekly Report (MMWR)^a^ Week; National Center for Health Statistics, United States, 2015-16 through 2025-26 influenza seasons

^a^MMWR weeks are defined by the Saturday week-ending date. For influenza surveillance, each season begins in MMWR Week 40, the first week of October, and ends in MMWR Week 39, the last week of September of the following year

**Supplementary Figure 8**: Reported influenza-associated pediatric deaths, 2010-11 through 2025-26 influenza seasons, reported as of October 28, 2026, United States, September 28, 2025- October 3, 2026 (Morbidity and Mortality Weekly Report (MMWR)^a^ Weeks 40-39)

Number of influenza-associated deaths

| Season | Total |
| --- | --- |
| 2010-11 | 124 |
| 2011-12 | 37 |
| 2012-13 | 169 |
| 2013-14 | 109 |
| 2014-15 | 147 |
| 2015-16 | 95 |
| 2016-17 | 110 |
| 2017-18 | 188 |
| 2018-19 | 145 |
| 2019-20 | 199 |
| 2020-21 | 1 |
| 2021-22 | 49 |
| 2022-23 | 187 |
| 2023-24 | 211 |
| 2024-25 | 298 |
| 2025-26 | 198 |

^a^MMWR weeks are defined by the Saturday week-ending date. For influenza surveillance, each season begins in MMWR Week 40, the first week of October, and ends in MMWR Week 39, the last week of September of the following year

**Supplementary Table 3a:** Preliminary Influenza Disease Burden Estimates by Age Group, United States 2025-26 influenza season

|  | **Symptomatic Illnesses** | | **Medical Visits** | | **Hospitalizations** | | **Deaths** | |
| --- | --- | --- | --- | --- | --- | --- | --- | --- |
| **Age group** | **Estimate** | **95% UI†** | **Estimate** | **95% UI†** | **Estimate** | **95% UI†** | **Estimate** | **95% UI†** |
| **0-4 years** | 3,100,000 | (2,600,000, 6,200,000) | 2,100,000 | (1,600,000, 4,200,000) | 21,000 | (18,000, 43,000) | 33 | (0, 94) |
| **5-17 years** | 10,000,000 | (9,000,000, 21,000,000) | 5,400,000 | (4,100,000, 11,000,000) | 29,000 | (25,000, 58,000) | 160 | (55, 380) |
| **18-49 years** | 12,000,000 | (9,700,000, 25,000,000) | 4,600,000 | (3,100,000, 9,400,000) | 69,000 | (55,000, 140,000) | 1,400 | (1,100, 3,200) |
| **50-64 years** | 5,900,000 | (5,000,000, 12,000,000) | 2,500,000 | (1,700,000, 5,400,000) | 63,000 | (53,000, 130,000) | 3,800 | (2,800, 7,500) |
| **65+ years** | 2,800,000 | (2,100,000, 5,400,000) | 1,500,000 | (1,000,000, 3,200,000) | 250,000 | (190,000, 490,000) | 16,000 | (15,000, 42,000) |
| **All ages** | 35,000,000 | (30,000,000, 49,000,000) | 16,000,000 | (14,000,000, 23,000,000) | 430,000 | (360,000, 690,000) | 22,000 | (20,000, 48,000) |

**Supplementary Table 3b**: Preliminary Influenza Prevented Disease Burden Estimates by Age Group, United States 2025-26 influenza season

|  | **Symptomatic Illnesses** | | **Medical Visits** | | **Hospitalizations** | | **Deaths** | |
| --- | --- | --- | --- | --- | --- | --- | --- | --- |
| **Age group** | **Estimate** | **95% UI†** | **Estimate** | **95% UI†** | **Estimate** | **95% UI†** | **Estimate** | **95% UI†** |
| **0-4 years** | 780,000 | (490,000, 1,600,000) | 530,000 | (330,000, 1,100,000) | 3,900 | (1,800, 7,900) | 6 | (0, 18) |
| **5-17 years** | 1,500,000 | (890,000, 3,100,000) | 790,000 | (460,000, 1,600,000) | 5,100 | (3,000, 11,000) | 29 | (8, 77) |
| **18-49 years** | 1,100,000 | (760,000, 2,400,000) | 420,000 | (280,000, 880,000) | 6,600 | (4,600, 14,000) | 130 | (86, 320) |
| **50-64 years** | 700,000 | (520,000, 1,400,000) | 300,000 | (220,000, 610,000) | 5,600 | (4,000, 11,000) | 340 | (210, 660) |
| **65+ years** | 540,000 | (370,000, 1,100,000) | 300,000 | (210,000, 620,000) | 55,000 | (40,000, 110,000) | 3,600 | (3,000, 9,400) |
| **All ages** | 4,700,000 | (3,800,000, 6,800,000) | 2,300,000 | (1,900,000, 3,400,000) | 76,000 | (60,000, 130,000) | 4,100 | (3,600, 10,000) |
